# Dobutamine Stress Echocardiography in Low-Gradient Aortic Stenosis

**DOI:** 10.1101/2023.02.27.23286540

**Authors:** Nils Sofus Borg Mogensen, Mulham Ali, Rasmus Carter-Storch, Mohamed-Salah Annabi, Jasmine Grenier-Delaney, Jacob Eifer Møller, Kristian Altern Øvrehus, Patricia A Pellikka, Philippe Pibarot, Marie-Annick Clavel, Jordi Sanchez Dahl

## Abstract

**Background:** Dobutamine stress echocardiography (DSE) is recommended by guidelines to distinguish between true-severe and pseudo-severe aortic stenosis (AS) in patients with low-gradients and left ventricular ejection fraction (LVEF) <50%. However, DSE has mostly been tested in the setting of LVEF<35% and determination of AS severity has mostly been based on outcome data and surgeon’s evaluation. The purpose of this study was to examine the diagnostic accuracy of guideline recommendations for DSE, in patents with low-gradient severe AS with a wide range of LVEF and to examine the interaction between the diagnostic accuracy of DSE and LVEF. Furthermore, we wanted to study the safety and feasibility of DSE in patients with LVEF>50%.

**Methods:** Patients with aortic mean gradient <40 mmHg, AVA <1.0 cm^2^, and stroke volume index ≤35 mL/m^2^ undergoing DSE and Cardiac Computer Tomography (C-CT) were identified from three prospectively collected patient cohorts, and stratified according to LVEF; LVEF <35%, LVEF 35-50% & LVEF >50%. Severe AS was defined as AVC score ≥2000 AU among men, and ≥1200 AU for women on C-CT.

**Results:** Two hundred twenty-one patients were included in the study. Seventy-eight (35%) presented with LVEF <35%, 67 (30%) with LVEF 35-50%, and 76 (34%) with LVEF >50%. DSE was performed without adverse symptoms or significant arrhythmias in 215 (96%) patients and stroke volume increased uniformly with no significant differences between groups (p=0.28).

Mean gradient and V_max_ during DSE showed significantly diagnostic heterogeneity between LVEF groups, being most precise when LVEF <35% (both AUC=0.90), albeit with optimal thresholds of 30 mmHg & 377 cm/s, and a limited diagnostic yield in patients with LVEF≥35% (AUC=0.67 in LVEF 35-50% and AUC 0.65 in LVEF≥35%). Using guideline thresholds led to a sensitivity and specificity of 49%/84% for all patients with LVEF <50%.

**Conclusion:** While DSE is safe and leads to a uniform increase in stroke volume in patients with low gradient AS regardless of baseline LVEF, the association between DSE gradients and AS severity assessed by C-CT demonstrates important heterogeneity depending on LVEF, with highest accuracy in patients with LVEF <35%.

**Clinical perspective:** *What is new?:*

- Dobutamine stress echocardiography (DSE) is safe in patients with low-gradient AS with LVEF >50%, and leads to similar increase in stroke volume as in patients with LVEF <50%.
- The diagnostic accuracy of DSE, compared to AVC as the reference for severe AS, depends on LVEF with highest accuracy in patients with LVEF <35%.
- Suggested reference thresholds for DSE may not be the most accurate for AS severity, when compared to AVC.

*What are the clinical implications?:*

- Based on our study, we suggest that DSE should primarily be used for determining AS severity in patients with LVEF <35%.

---

While the diagnosis of severe aortic stenosis (AS) is straightforward when aortic valve area (AVA) is <1.0 cm^2^ and transvalvular mean gradient is ≥40 mmHg,^1^ diagnosis may be more challenging when gradients are low. When this occurs in the presence of reduced stroke volume (SV) and left ventricular ejection fraction (LVEF)<50%, guidelines recommend the use of dobutamine stress echocardiography (DSE) to distinguish between true-severe –and pseudo-severe AS.^2-4^ However, only few data support a LVEF threshold of 50% as most studies have tested DSE in patients with severely reduced LVEF. Furthermore, evidence suggests that in AS LVEF may even be considered reduced already when less than 60%,^2, 5^ explaining why some, advocate DSE may provide diagnostic information even when LVEF >50%.^6^ However, all these studies are limited by the lack of a clear gold-standard of assessing AS severity,^7, 8^ and have thus largely been based on prognostic data^9^ rather than objective measures for AS severity per se. This poses a potential problem as DSE findings not being blinded for the clinicians, may have influenced decision making, potentially biasing the clinical end-point,^10-12^ even more so as even moderate AS may be associated with a poor prognosis when LVEF is reduced.^13^

Aortic valve calcification (AVC) assessed by cardiac computer tomography (C-CT) has recently emerged as an additional method of determining AS-severity.^3^ AVC has been demonstrated to clearly discriminate between moderate and severe AS,^14^ and is associated with outcome.^15^ Accordingly, ESC and AHA/ACC recommend the use of AVC to diagnose severe AS, in particular among patients with low-gradient AS.^2, 3^ The purpose of this study was to examine the diagnostic accuracy of guideline recommendations for DSE in low-gradient AS, in patents with a wide range of LVEF and to examine if an interaction between the diagnostic accuracy of DSE and LVEF exist. Furthermore, we studied the safety and feasibility of DSE in patients with LVEF>50%.

## Methods

We identified patients aged ≥18 years with low-gradient AS (aortic mean-gradient<40 mmHg and AVA<1.0 cm^2^) and stroke volume index (SV_i_) ≤35 mL/m^2^ from two prospectively collected cohorts at Quebec Heart and Lung Institute, Canada^12, 16^ and a prospective cohort collected at Odense University Hospital, Denmark between 2019 and 2022.

For this study, we excluded patients with missing DSE or C-CT data or with concomitant moderate or severe valvular heart disease other than AS. Patients were stratified in three subgroups according to LVEF (LVEF <35%, LVEF 35-50% and LVEF >50%) at the baseline evaluation which included a clinical examination, transthoracic echocardiography and DSE.

Research was performed in accordance with the Declaration of Helsinki, and informed consent was obtained according to approval by each institutional review board.

### Echocardiography

Patients underwent a comprehensive transthoracic echocardiographic examination using commercially available ultrasound systems in accordance with the American Society of Echocardiography guidelines^17, 18^ with interpretation performed by experienced cardiologists. Doppler values were calculated as the average of three cardiac cycles for patients with sinus rhythm and five cycles for atrial fibrillation. LV outflow tract diameter was measured in the parasternal long-axis view in early systole from the point of aortic cusp insertion into the interventricular septum to the point of aortic cusp insertion into the intervalvular fibrosis. AVA was estimated by quantitative Doppler ultrasound using the continuity equation. Peak and mean flow velocity across the valve were determined in the window where the highest velocity could be recorded using continuous wave Doppler with the cursor as parallel as possible with the flow across the valve. Peak and mean transvalvular gradients were estimated using the modified Bernoulli equation.^1^ LVEF was determined by the Simpson biplane method. LV stroke volume was calculated using pulsed-wave Doppler as the product of the LV outflow area and LV outflow tract time velocity integral and indexed for body surface area (SVi). LV mass index was estimated using the Devereux formula.^19^ In men LV mass index >116 g/m^2^ and in women, >104 g/m^2^ were considered indicative of LV hypertrophy.^20^ Relative wall thickness (RWT) was calculated for assessment of LV geometry using the formula 2xposterior wall thickness/LV internal diameter in diastole. Increased relative wall thickness was present when this ratio was>0.42.^21^ Normal geometry was present when LV mass index and RWT were normal, concentric remodeling with increased RWT and normal LV mass index, eccentric LV hypertrophy with increased LV mass index and normal RWT, and concentric LV hypertrophy when both were increased.

Valvulo-arterial impedance was calculated using the formula (systolic blood pressure + mean aortic valve pressure gradient)/SVi.^22^ Pulse pressure was measured as the difference between systolic and diastolic blood pressure, and systemic arterial compliance was calculated as SVI/pulse pressure,^22^ Systemic vascular resistance was calculated as 80*(1/3*systolic blood pressure+2/3*diastolic blood pressure)/cardiac output.^22^

### Dobutamine stress echocardiography

A comprehensive DSE was performed in all patients. Dobutamine infusion was initiated at a dose of 5 µg/kg/min and increased every 3rd min to a maximal dosage of 20µg/kg/min. DSE was terminated early if an adverse event occurred, if the patient became symptomatic during the examination, or if a mean gradient > 40 mmHg was recorded in patients with LVEF <50%. Contractile reserve was defined as an increase in SV_i_ exceeding 20%. We defined patients as still having a low-flow state if SV_i_ was ≤35 mL/m^2^ despite DSE.

Three cardiac cycles were averaged for all measurements in patients with sinus rhythm, and five cardiac cycles were averaged for patients with atrial fibrillation. Echocardiographic measurements of aortic flow and LV were obtained at each stage. During DSE heart rate, systolic and diastolic blood pressure was recorded at each dose increment.

### Cardiac computer tomography

All patients underwent a non-contrast C-CT with a tube potential at 120 kV. Operators blinded to patient clinical and echocardiographic data performed all MDCT analyses. The aortic valve was visualised in multiple planes, and careful measurement section by section aimed to accurately distinguish contiguous calcium in coronary arteries, mitral valve annulus or aortic wall. AVC score was assessed using the Agatston method, and expressed in arbitrary units. We defined severe AS using the sex-specific thresholds recommended by guidelines, AVC score ≥2000 AU in males, and ≥1200 AU in females on C-CT.^2, 3, 23^

### DSE safety end-points

Patients undergoing DSE were safety monitored for adverse events. Adverse events during DSE were defined as new onset of complex ventricular arrhythmia, a rise in systolic blood pressure ≥200mmHg, a decrease in systolic blood pressure <80 mmHg, LV outflow tract peak-flow velocity ≥ 2.0 m/sec or systolic anterior motion of the mitral valve.

### Statistical analysis

Continuous variables were tested for normality with the Shapiro-Wilk test and are expressed as either mean ± SD or median and interquartile range. Differences in values between groups were tested by 1-way ANOVA. Categorical variables are expressed as number and percentages, and tested by the χ^2^ exact test of Fisher exact test when the expected value in any of the cells of a contingency table was below 5.

Time between C-CT and DSE, and AVC are presented as median and interquartile range, and differences were tested using the Kruskal-Wallis test because of non-Gaussian distribution for these variables. Correlations were obtained using Spearman rank test. For overall tests, a P value of <0.05 were considered significant and 2-sided tests were used. Comparison of each method’s predictive capability was performed by comparing the C-statistic derived from the area under the receiver operating characteristic curves using the generalized U-statistic as proposed by DeLong et al.^24^ Statistical analysis was performed with STATA/SE V.17.0 (StataCorp, Texas, USA) software.

## Results

We identified 635 patients with low-gradient AS, and excluded those with missing C-CT data (n=240), missing DSE data (n=178), and AVA>1.0 cm^2^ (n=8) or mean gradient>40 mmHg (n=1) leaving 221 patients (n=150 Quebec Heart Institute, n=71 Odense University Hospital) in this study. Seventy-eight (35%) presented with LVEF <35%, 67(30%) with LVEF 35-50%, and 76(34%) with LVEF >50%. DSE and C-CT were performed within a median timespan of 12 [1; 26] days, with no difference between groups (p=0.17).

Patients with LVEF >50% were more likely to be women, had less symptoms, and were less likely to have a pacemaker or ICD-device (Table 1). Severe AS, as evaluated by AVC, was present among 102 (46%) patients with no differences between groups, (55% vs 37% vs 46%, LVEF<35% LVEF 35-50%, LVEF>50% respectively, p = 0.10). Patients with LVEF >50% presented with higher V_max_ (304±45 vs 302±43vs 324±37cm/s, p<0.01) and mean gradient (22±7 vs 23±7 vs 25±6mmHg, p=0.03), despite similar AVA.

**Table 1:**
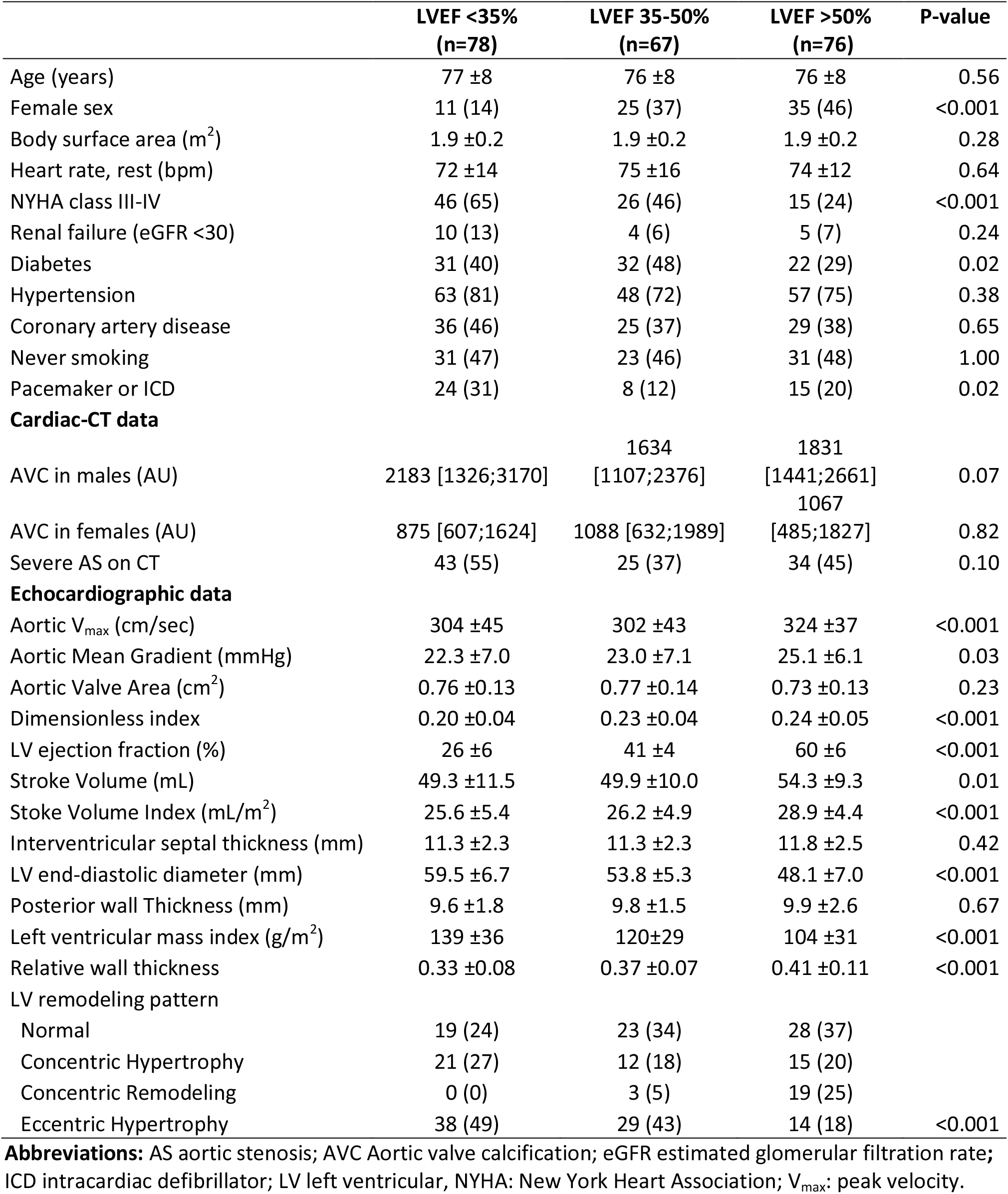
Baseline Characteristics.

An inverse relationship was seen between LVEF and left ventricular diameter (r=-0.65, p<0.001) (table 1). Patients with LVEF<35% had higher LVM_i_ and lower RWT than patients with LVEF>35%. These differences translated to significant differences in geometric patterns with eccentric hypertrophy being the most common pattern among patients with LVEF<35%, and patients with LVEF 35-50%, and normal geometry in patients with LVEF>50% (p<0.001) (Table 1). All patients had SV_i_<35 ml/m^2^, but a higher resting SV_i_ was present in patients with LVEF >50% (26±5vs 26±5 vs 29±4mL/m^2^, p<0.01, Table 1).

### Dobutamine stress echocardiography

DSE was performed without adverse symptoms in two-hundred-fifteen (97%) patients. However, in six patients DSE was discontinued prematurely due to adverse symptoms (n=1 systolic blood pressure>200 mmHg, n=1 angina, n=1 dyspnea=1, n=3 other discomfort) being most common among patients with LVEF>50% (0% vs 2% vs. 9%, p=0.04, LVEF<35%; LVEF 35-50% and LVEF>50% respectively). In contrast no ventricular arrhythmia during DSE were seen in patients with LVEF>50% while 8 patients with LVEF<50% (4 in LVEF<35% and 4 in LVEF 35-50%) experienced complex ventricular arrhythmia during DSE (Table 2).

**Table 2:** Safety parameters during dobutamine stress echocardiography.

|  | LVEF <35%<br>(n=78) | LVEF 35-50%<br>(n=67) | LVEF >50%<br>(n=76) | P-value |
| --- | --- | --- | --- | --- |
| Max systolic BP during DSE (mmHg) | 133 ±27 | 141 ±21 | 149 ±23 | <0.001 |
| Systolic BP during max DSE dose (mmHg) | 130 ±27 | 134 ±21 | 142 ±23 | 0.03 |
| Systolic BP <80mmHg | 0 (0) | 0 (0) | 1 (1) | 0.38 |
| Systolic BP >200mmHg | 1 (1) | 0 (0) | 2 (3) | 0.40 |
| DSE stopped because of adverse symptoms | 0 (.0) | 1 (2) | 5 (7) | 0.03 |
| Chest pain | 1 (1) | 2 (3) | 2 (3) | 0.76 |
| Ischemia on ECG | 0 (0) | 0 (0) | 0 (0) | 0.99 |
| Newly onset SVT | 1 (1) | 1 (2) | 1 (1) | 0.99 |
| Complex ventricular arrhythmia | 4 (5) | 4 (6) | 0 (0) | 0.11 |

**Abbreviations:** BP blood pressure; DSE dobutamine stress echocardiography; ECG electrocardiogram; SVT supraventricular tachycardia.

During DSE, stroke volume increased uniformly with no between groups differences (15±12mL vs 13±13mL vs 12±12mL, p=0.28), however more patients with reduced LVEF increased their SV_i_ ≥20% (65% vs 49% vs 44%, p=0.03). One-hundred-twenty (56%) patients still had SV_i_ ≤35 mL/m^2^ at the end of DSE evenly split between LVEF subgroups (Table 3). Stroke volume increased unanimously regardless of a history of coronary artery disease or intracardiac device, beta-blocker therapy or AS severity (Supplemental figure 1). Despite similar systemic vascular resistance and valvulo-arterial impedance during baseline and dobutamine infusion, patients with LVEF>35% experienced lower systemic arterial compliance than those with LVEF<35% both at baseline and during DSE (0.56±0.21 vs 0.48±0.17 vs 0.50±0.14ml/m^2^/mmHg, p=0.02, LVEF<35%, LVEF 35-50%, LVEF>50% respectively).

**Table 3:** Dobutamine stress echocardiograhic parameters.

|  | LVEF <35%<br>(n=78) | LVEF 35-50%<br>(n=67) | LVEF >50%<br>(n=76) | P-value |
| --- | --- | --- | --- | --- |
| **Hemodynamic data** |  |  |  |  |
| Systolic BP, rest (mmHg) | 123 ±21 | 129 ±18 | 137 ±21 | <0.001 |
| Diastolic BP, rest (mmHg) | 72 ±12 | 71 ±10 | 75 ±13 | 0.08 |
| Systolic BP during max DSE dose (mmHg) | 130 ±27 | 134 ±21 | 142 ±23 | 0.03 |
| Diastolic BP during max DSE dose (mmHg) | 67 ±14 | 67 ±13 | 69 ±12 | 0.68 |
| BP pressure change (mmHg) | 10 ±23 | 9 ±22 | 9 ±20 | 0.97 |
| Max heart rate during DSE (bpm) | 93 ±18 | 103 ±25 | 103 ±24 | 0.08 |
| Change in heart rate (bpm) | 25 ±20 | 21 ±15 | 24 ±19 | 0.78 |
| Zva, rest (mmHg/mL/m<sup>2</sup>) | 5.85 ±1.47 | 6.00 ±1.27 | 5.76 ±1.17 | 0.55 |
| Zva, DSE (mmHg/mL/m<sup>2</sup>) | 5.01 ±1.41 | 5.48 ±1.45 | 5.38 ±1.28 | 0.24 |
| Zva, diff. (mmHg/mL/m<sup>2</sup>) | 1.12 ±0.95 | 1.10 ±0.73 | 0.93 ±0.63 | 0.39 |
| Systemic Arterial Compliance, rest (Units) | 0.56 ±0.21 | 0.48 ±0.17 | 0.50 ±0.14 | 0.02 |
| Systemic Arterial Compliance, DSE (Units) | 0.60 ±0.21 | 0.50 ±0.17 | 0.51 ±0.18 | 0.02 |
| Systemic Arterial Compliance, diff. (Units) | 0.17 ±0.11 | 0.12 ±0.12 | 0.11 ±0.10 | 0.02 |
| SVR, rest (mmHg/L/min) | 1239 ±451 | 1241 ±395 | 1221 ±380 | 0.97 |
| SVR, DSE (mmHg/L/min) | 1209 ±562 | 1269 ±389 | 1228 ±375 | 0.85 |
| SVR, diff. (mmHg/L/min) | 173 ±177 | 199 ±164 | 144 ±131 | 0.28 |
| Stroke Volume, DSE (mL) | 64.6 ±15.5 | 63.3 ±16.4 | 65.6 ±15.6 | 0.68 |
| Low-Flow(<35ml/m<sup>2</sup>) despite DSE | 42 (58) | 41 (62) | 37 (49) | 0.29 |
| Change in SV (mL) | 14.7 ±11.5 | 13.1 ±12.5 | 11.5 ±11.8 | 0.28 |
| Change in SVi >20% | 47 (65) | 32 (49) | 33 (44) | 0.03 |
| Aortic V<sub>max</sub>, DSE (cm/sec) | 377 ±55 | 378 ±58 | 405 ±47 | <0.001 |
| Change in V<sub>max</sub> (cm/sec) | 70 ±35 | 76 ±39 | 82 ±33 | 0.14 |
| Aortic Mean Gradient, DSE (mmHg) | 32.7 ±10.1 | 34.2 ±11.5 | 38.7 ±9.9 | <0.001 |
| Change in Mean Gradient (mmHg) | 10.6 ±5.7 | 11.6 ±6.7 | 13.7 ±6.5 | 0.01 |
| Aortic Valve Area, DSE (cm<sup>2</sup>) | 0.94 ±0.28 | 0.97 ±0.33 | 0.91 ±0.21 | 0.52 |
| Change in AVA (cm<sup>2</sup>) | 0.19 ±0.23 | 0.21 ±0.30 | 0.19 ±0.15 | 0.86 |
| DSE Dimensionless index | 0.24 ±0.05 | 0.28 ±0.06 | 0.30 ±0.07 | <0.001 |

**Abbreviations:** AVA aortic valve area BP blood pressure; DSE dobutamine stress echocardiography; SV stroke volume; SV<sub>i</sub> stroke volume indexed; SVR Systemic Vascular Resistance; V<sub>max</sub> peak velocity; Zva Valvulo-arterial Impedance .

### Classifying severe AS in the entire cohort

Receiver operating characteristic curves for DSE-derived mean gradient, V_max_ and AVA are provided in Figure 2. C-statistics for the mean gradient (AUC=0.73) were non-significant higher than V_max_ (AUC=0.70) but significantly higher than AVA (AUC=0.61) (p=0.01, Figure 2), with differences remaining non-significant in a direct comparison between mean gradient and V_max_ after including AVA in both models (AUC 0.74 vs 0.72, p=0.12).

**Figure 1:**
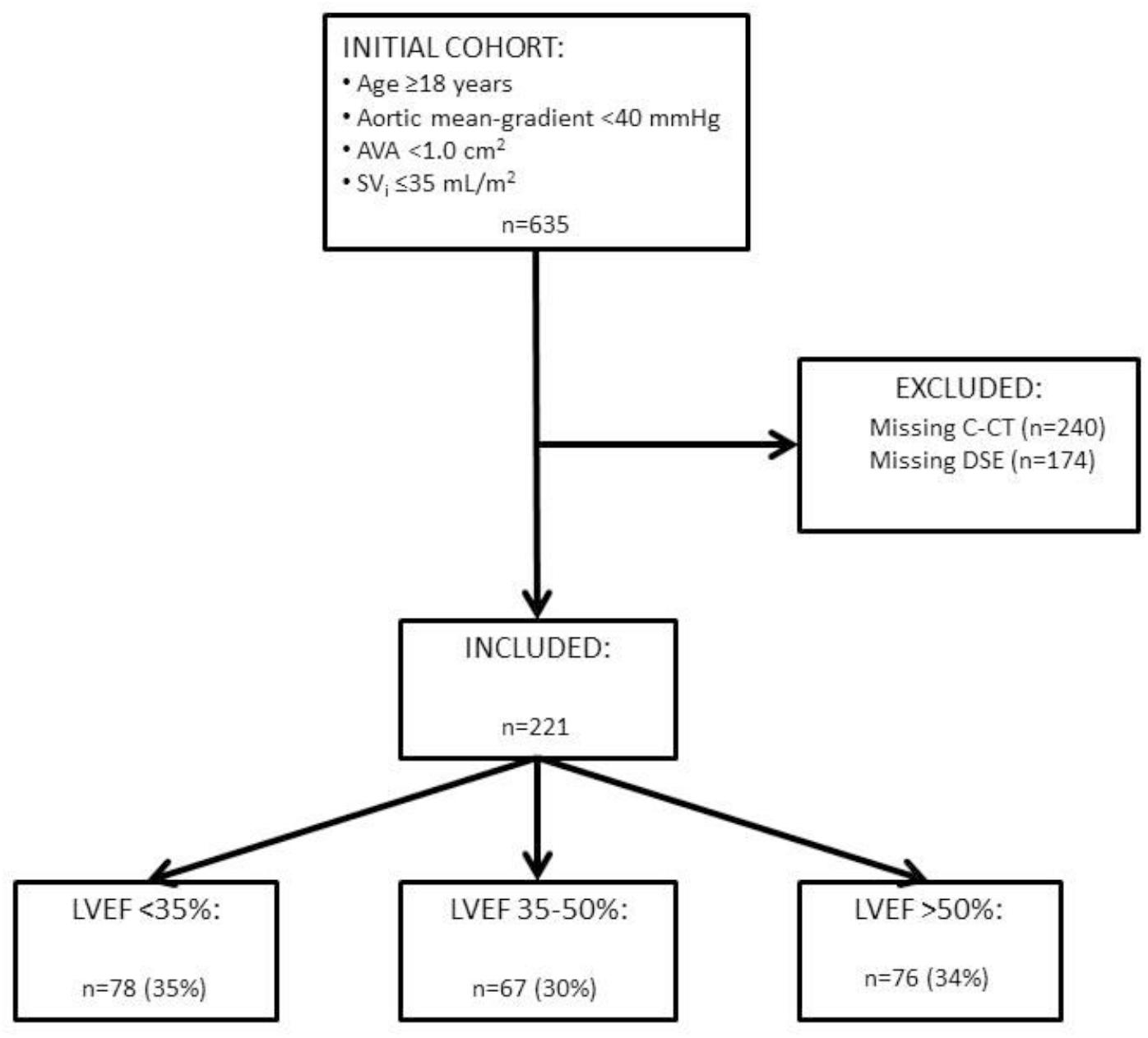
Consort flowchart of patient inclusion. Consort Flowchart showing the process of inclusion and exclusion of patients. AS: aortic stenosis, AVA: aortic valve area, C-CT: Cardiac Computer tomography, DSE: dobutamine stress echocardiography, LVEF: left ventricular ejection fraction SV_i_: stoke volume indexed.

**Figure 2:**
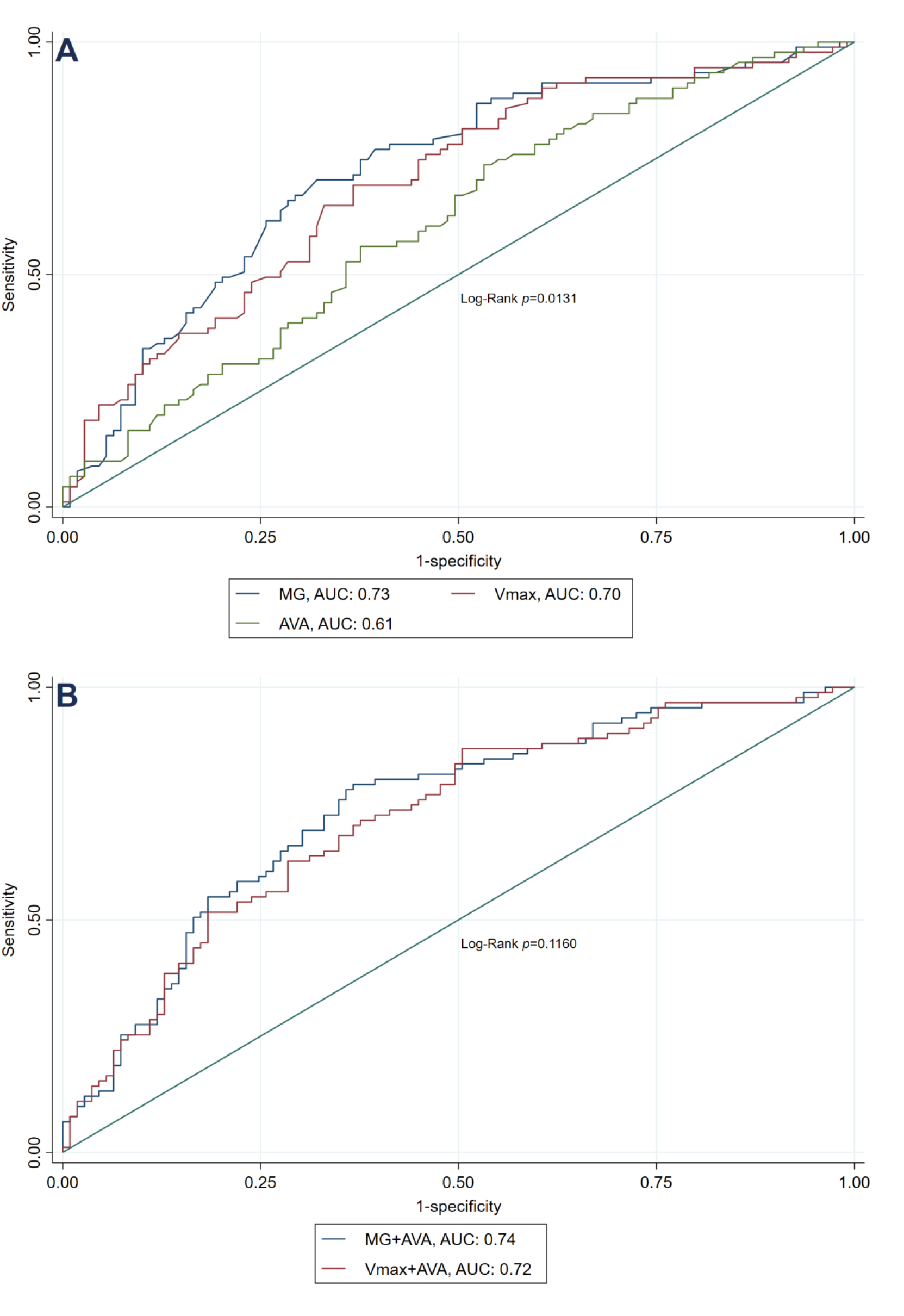
Receiver-Operating Characteristic to identify severe AS in the entire cohort. Receiver-Operating Characteristic to identify severe aortic stenosis (AS) in the entire cohort of 221 patients with low-gradient AS (mean gradient <40 mmHg and aortic valve area <1.0 cm^2^) and stroke volume index ≤35 mL/m^2^ according to (**A**) mean gradient, peak velocity and aortic valve area, (**B**) Combination of mean gradient + aortic valve area and peak velocity + aortic valve area against computed tomography aortic valve calcification. MG: mean gradient, V_max_: peak velocity, AVA: aortic valve area, AUC: area under the curve, DSE: Dobutamine Stress Echocardiography.

The optimal cut-off points for discriminating between severe and pseudo-severe-AS during DSE was identified from ROC-curves; 34 mmHg, 389 cm/s and 0.9 cm^2^ with a sensitivity and specificity of 75%/64% vs 70%/63% vs 63%/55% when utilising these thresholds (mean gradient, V_max_ and AVA respectively) (Table 4). Applying either of the guideline specific thresholds; mean gradient > 40mmHg, V_max_ > 400cm/s or AVA <1.0cm^2^, lead to a sensitivity and specificity of 49%/78% vs 64%/66%, vs 41%/78%, respectively (Table 4). Combining guideline recommendations for AVA and either mean gradient or V_max_ led to a sensitivity and specificity of 54%/77% for the entire cohort.

**Table 4:** Diagnostic accuracy of dobutamine stress echocardiography in differentiating severe from pseudo-severe AS.

| Optimal cut-off points |  |  |  | Guideline recommendations |  |  |  |
| --- | --- | --- | --- | --- | --- | --- | --- |
| Marker | Sensitivity | Specificity | CC | Marker | Sensitivity | Specificity | CC |
| MG<br>(≥34mmHg) | 75% | 64% | 69% | MG<br>(≥40mmHg) | 49% | 78% | 65% |
| V<sub>max</sub><br>(≥389cm/s) | 70% | 63% | 66% | V<sub>max</sub><br>(≥400cm/s) | 64% | 66% | 65% |
| AVA<br>(<0.9cm<sup>2</sup>) | 63% | 55% | 60% | AVA<br>(<1.0cm<sup>2</sup>) | 41% | 78% | 58% |
| Combined<br>(for LVEF<br><50%) | 48% | 87% | 67% | Combined<br>(for LVEF<br><50%) | 50% | 67% | 67% |

**Abbreviations:** AVA aortic valve area; CC correct classification; MG mean gradient.

### Classifying severe AS according to LVEF subgroups

Comparing the ability of DSE variables to predict severe AS demonstrated significant heterogeneity between LVEF subgroups. AVA displayed a rather uniform optimal cut-off between LVEF subgroups (1.0 cm^2^ vs 0.9 cm^2^ vs 0.8 cm^2^, LVEF<35%, LVEF 35-50%, LVEF>50%, respectively) with similar c-statistics (AUC=0.68, vs AUC 0.62 vs 0.54, p=0.36, LVEF<35, LVEF35-50%, LVEF>50% respectively, Figure 3). In contrast, the association of both mean gradient and V_max_ with AS severity was more accurate in the LVEF<35% group (Mean gradient; AUC=0.90 vs.0.67 vs 0.65, p=0.0007, V_max_; AUC=0.90 vs 0.66 vs 0.60, p=0.0001, Figure 3), with different optimal cut-off points (30 vs 45 vs 37mmHg & 377 vs 430 vs 400cm/s, LVEF<35%, LVEF 35-50%, LVEF>50%, respectively).

**Figure 3:**
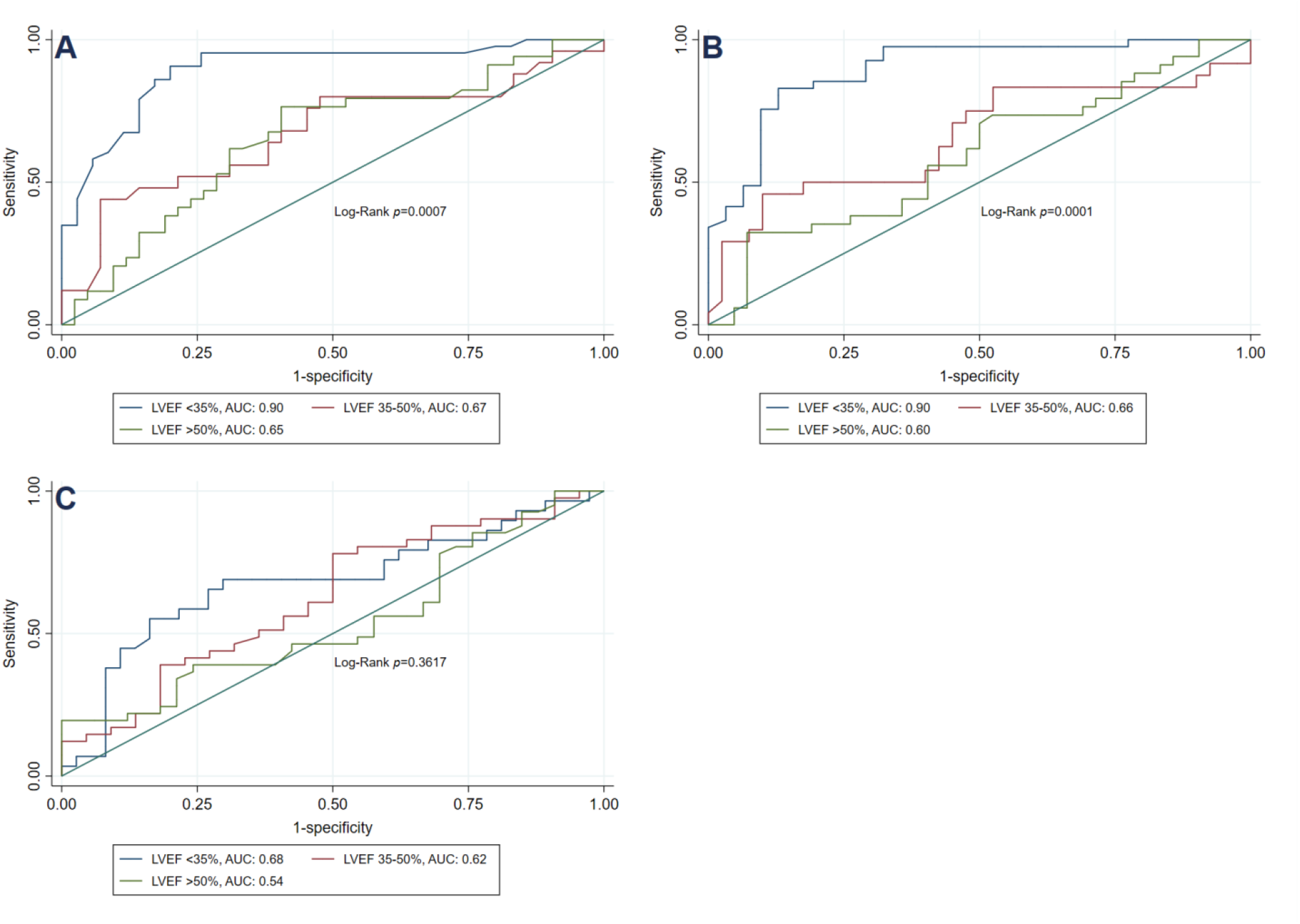
Receiver-Operating Characteristic to identify severe AS in LVEF subgroups. Receiver-Operating Characteristic to identify severe aortic stenosis (AS) in LVEF subgroups; LVEF <35%, 35-50% and >50% respectively, according to (**A**) mean gradient, (**B**) peak velocity, and (**C**) aortic valve area against computed tomography aortic valve calcification. LVEF: left ventricular ejection fraction, AUC: area under the curve.

Using guideline thresholds for both AVA and either mean gradient or V_max_ led to a sensitivity and specificity of 54%/93% vs 41%/78% vs 64%/63%, LVEF<35%, LVEF 35-50%, LVEF>50%, respectively (Figure 4). For all patients with LVEF <50%, the sensitivity and specificity was 49%/84%.

**Figure 4:**
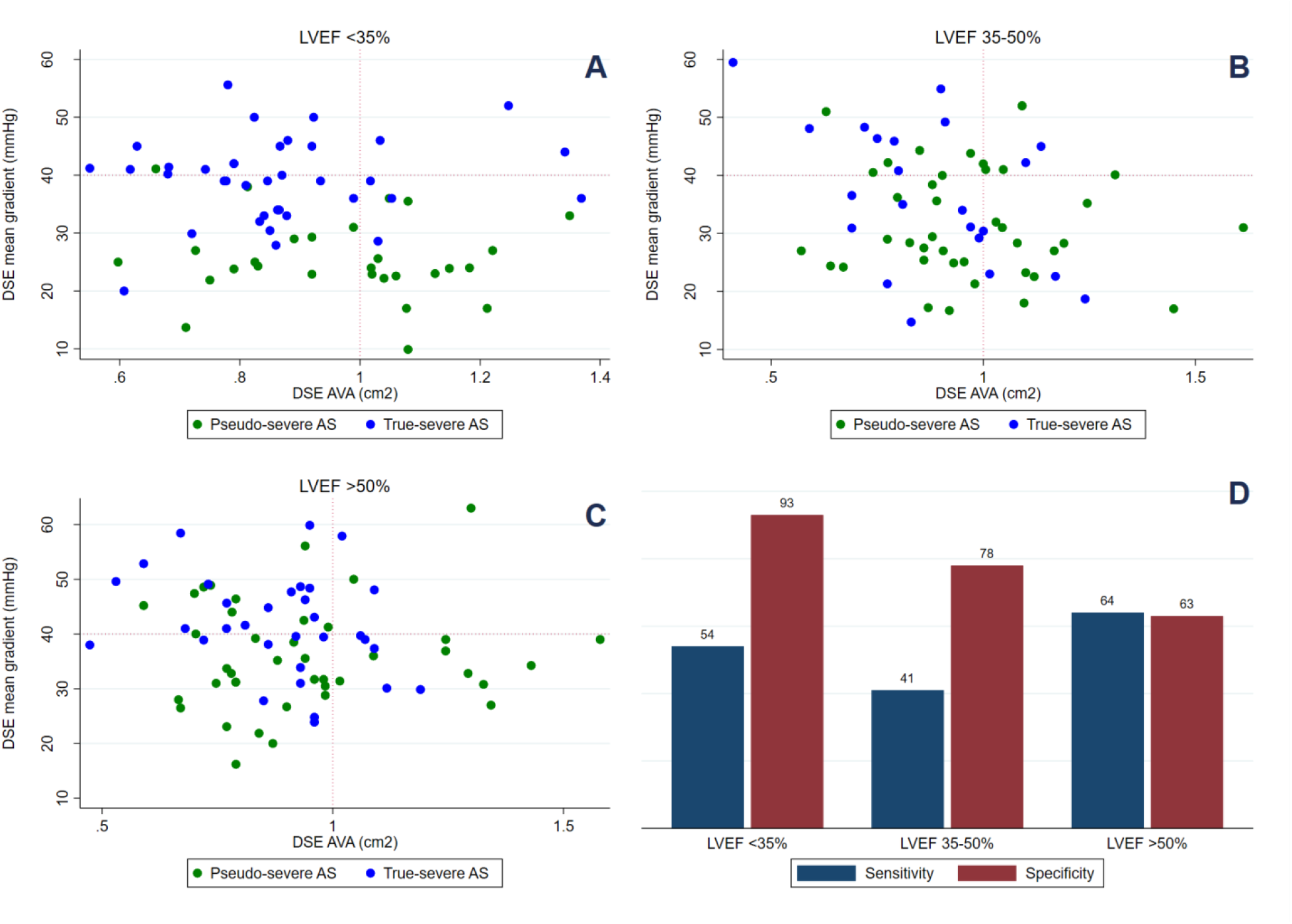
Pseudo-severe and True-severe AS in LVEF subgroups. Scatterplots for LVEF subgroups; LVEF <35%, 35-50% and >50% respectively. Green represents Pseudo-severe aortic stenosis (AS) and Blue True-severe AS determined by aortic valve calcification. Dashed lines represents guideline cut-offs for mean gradient (40 mmHg) and aortic valve area (>1.0cm^2^); (**A**) LVEF <35%, (**B**) LVEF 35-50%, (**C**) LVEF >50%. (**D**) Sensitivity and specificity for severe AS according to guideline cut-offs for DSE for each LVEF subgroups; LVEF <35%, 35-50% and >50% respectively. LVEF: left ventricular ejection fraction, DSE: dobutamine stress echocardiography, AVA: aortic valve area, AS: aortic stenosis.

Optimal cutoff points were similar between LVEF subgroups, regardless of including center or flow-state during DSE.

## Discussion

In this study with prospectively enrolled patients with low-gradient AS undergoing DSE we demonstrate four novel findings. 1) DSE is safe with few patients experiencing dobutamine associated complications in patients with reduced as well as preserved LVEF. 2) DSE leads to a uniform increase in stroke volume in patients with low gradient aortic stenosis regardless of baseline LVEF. 3) The transvalvular mean gradient and transvalvular peak-velocity during DSE outperformed AVA in diagnosing severe AS adjudicated with C-CT aortic valve calcium score. However, mean gradient was associated with a lower sensitivity but higher specificity than V_max_. Utilizing the guideline recommendations of combining transvalvular gradients with AVA resulted in a specific but non-sensitive discrimination between severe and pseudo-severe AS in patients with LVEF<50%, highlighting that a large proportion of patients with high AVC are labeled as pseudo-severe AS based on DSE findings. This suggests that there is a discrepancy between the guideline recommended thresholds of DSE and CT indices of severe AS. 4) Although AVA during DSE provided modest, but similar information regardless of LVEF, with a rather uniform optimal cut-off of 0.9 cm^2^, both transvalvular mean-gradient and peak-velocity demonstrated important heterogeneity with outstanding discrimination in patients with LVEF<35% while only modest in those with LVEF>35%. In addition, the optimal discriminatory threshold was markedly different between LVEF groups with a mean-gradient 30 mmHg being the best cut-off in patients with LVEF<35%, and 40 mmHg in those with LVEF>35%. The later suggests that discrepancies exist between guidelines proposed thresholds for DSE and CT in the assessment of severe AS and reduced LVEF.

### LOW-GRADIENT AS AND DSE IN CURRENT GUIDELINES

While for decades it has been accepted that severe AS may occur despite low-gradients, when LVEF is reduced,^25^ it was not until the seminal paper by Hachicha and colleagues^26^ that it became clear that this could also be the case when LVEF>50%. Regardless of LVEF, low-gradient AS raises the diagnostic conundrum of differentiating between severe- and pseudo-severe AS.

Currently, guidelines distinguish between patients according to stroke volume. Patients with low-gradient AS under normal-flow conditions (SV_i_>35 ml/m^2^) are thought as having moderate AS., while further testing is required in patients with SV_i_<35 ml/m^2^to determine AS severity. For patients with LVEF<50%, guidelines recommend the use of DSE in order to increase flow and demonstrate high-gradients with a reduced AVA. ^2, 3^ This strategy has been recommended since deFilippi and colleagues suggested it in 1995,^27^ and Monin et al. in 2 subsequent larger populations tested the possible impact of DSE on outcome.^10, 11^ In the first study including forty-five patients with low-gradient AS and LVEF<30%, DSE with a mean dosage of 9 µg/kg/min lead to an increase in stroke volume, with 71% demonstrating contractile reserve with no adverse events. Despite similar baseline characteristics, patients with contractile reserve experienced better outcome, in particular if undergoing surgery.^10^ These findings were corroborated in a subsequent paper with a larger sample size including 136 patients with low-gradient AS patients and LVEF<35%.^11^

In line with these studies, we concordantly demonstrate that ∼2/3 of patients with LVEF<35% had contractile reserve, but as a novel finding show a lower rate of contractile reserve among patients with LVEF>35%. Although this could counterintuitively be interpreted as patients with higher baseline LVEF having a poorer ability to increase their contractile state, we speculate this rather reflects differences in LV-geometry, and baseline SV_i_ between groups. While reduced stroke volume in patients with LVEF<35% is the consequence of poor contractility, patients with LVEF>50% predominantly have a reduced stroke volume due to concentric LV remodeling that leads to small cavities and impeding normal stroke volume despite preserved LVEF.^28^ Accordingly, the combination of smaller cavities combined with a higher baseline SVi in patients with LVEF>50% challenges a SV_i_ increase > 20% as a criterion for contractile reserve. Thusly, caution should be taken when interpreting contractile reserve without regards to LVEF. This may pose a potential problem as guidelines recommend the use of DSE in patients with LVEF<50% while DSE, and contractile reserve, has mainly been studied in patients with LVEF<35%. ^7-12^

### C-CT RECOMMENDATIONS IN CURRENT GUIDELINES

A potential limitation of DSE is that AS severity may be challenging to grade when flow does not increase during DSE. In this setting, Cueff and colleagues demonstrated that C-CT derived AVC using a cut-off 1650 AU was able to distinguish severe from non-severe AS when LVEF<40%.^29^ Since, it has been demonstrated that the impact of calcification on AS severity is largely affected by sex, as females demonstrate a steeper association between calcium load and gradient than males and will experience severe AS despite less calcium.^30^ Accordingly, it has been demonstrated that AVC has different sex-specific thresholds for men and women for the same echocardiographic measurement of AS-severity.^23, 31^ Consequently, AVC has been implemented in the current ESC and AHA/ACC guidelines as a tool to differentiate between moderate and severe AS and is recommended in patients with low-gradient AS without contractile reserve on DSE or when LVEF>50%. It is thus interesting that among patients with LVEF<50% and severe AS ascertained by C-CT, 23% were labeled as having pseudo-severe AS when combining DSE gradients and AVA.

These discrepancies imply that a diagnosis of severe AS is more likely if we use C-CT assessed AVC rather than DSE data. Thus, patients with LVEF <35% and contractile reserve are less likely to be labeled as severe AS, based solely on DSE response, than patients without contractile reserve where determination of AS-severity relies on AVC.

As a consequence, it seems that while current DSE recommended thresholds are rather specific, they are also non-sensitive when compared to C-CT. This naturally raises the question of why the current thresholds have been chosen.

### THRESHOLDS IN CURRENT GUIDELINES

While most DSE studies in low-gradient AS have used the same thresholds for AS severity as in normal flow, confirmation of AS severity has solely been based on either patient outcome or surgeons’ evaluation during surgery as the indication of AS severity. ^10, 11^ In these studies, the decision of surgery was not blinded for DSE-response, and as no gold standard for severe AS existed outcome was chosen. The latter may pose a particular problem as it recently has become evident that even moderate AS may have an impact on outcome in patients with LVEF<50%.^13^

Indeed, a recent study by Annabi and colleagues intended to identify thresholds of severe AS based with DSE. However, confirmation was only possible in 87 of 186 patients, and only in 25 ascertained by CT.^32^ Thus, our study is the first to compare the response of DSE with AS severity ascertained by C-CT. Accordingly, it is interesting that while AVA demonstrated a rather uniform cut-off between LVEF-subgroups, both transvalvular mean gradient and transvalvular peak-velocity showed higher accuracy in patients with LVEF <35% with an optimal cut-off points of 30mmHg for transvalvular mean gradient and 377 cm/s for transvalvular peak-velocity.

These findings are in accordance with a previous paper by Nishimura and colleagues reporting that under DSE, an invasively measured gradient of <30 mmHg correlated with severe AS.^33^ In line with this, Annabi et al. found guideline DSE thresholds to have limited value in predicting stenosis severity, and demonstrated that lowering the cut-off point for transvalvular mean gradient to either 35 mmHg or 30 mmHg improved the diagnostic accuracy of DSE.^32^

A possible explanation to our finding could be that despite dobutamine raising stroke volume in most patients, almost half still experience a low-flow condition after dobutamine infusion suggesting that a lower gradient would be expected than during normal flow. Furthermore, while dobutamine has a positive inotropic effect on the heart stimulating *β*1 receptors, the effects on the *α* receptors may also alter vascular resistance, changing the ventriculo-arterial coupling and eventually lowering the transvalvular gradients.^34^ Suggestive of this, we demonstrated different systemic arterial compliance between LVEF groups.

Finally it is also possible that inconsistencies exist between gradients and AVA as a consequence of discrepancies also described in patients with normal systolic LV function. ^35^

## Study limitations

Assessment of AS severity is challenging as severity can be graded based on a wide range of variables with no clear gold standard. In this study, we used C-CT assessed AVC as the reference for adjudication of AS severity. Different concerns can been raised regarding this choice as 1) AVC does not quantify fibrotic tissue, which might play an important role in the development of low-gradient AS,^14, 30^ or in bicuspid valves; 2) the current thresholds are derived from patients with concordant AS and have not been validated in low-gradient AS patients, and 3) that reproducibility of AVC may be challenging. However, the Agatston method is a well-established marker of anatomic severity that has demonstrated to correlated with valve severity on explanted valves,^30^ AS hemodynamic severity measured by Doppler echocardiography,^29, 36^ and clinical outcomes.^15, 31^

Furthermore the reproducibility of AVC has been questioned. In our study, the measurement of AVC was done separately in each institution, but was standardized between centers and demonstrated excellent reproducibility.

While echocardiographic measurements were performed by experienced users, newer studies show, that in patients with atrial fibrillation the maximum peak velocity and mean gradient might be a more precise measurement for AS severity instead of an averaged peak velocity or mean gradient, when compared to C-CT.^37^

## Conclusion

The use of DSE is safe and leads to a uniform increase in stroke volume in patients with low gradient AS regardless of baseline LVEF. However, the association between DSE gradients and AS severity assessed by C-CT demonstrates important heterogeneity, with highest accuracy in patients with LVEF<35% but limited diagnostic yield when LVEF≥35%, and with different optimal diagnostic thresholds.

## Data Availability

not available

## Non-standard abbreviations and acronyms

AS: Aortic Stenosis
AUC: Area under curve
AVA: Aortic Valve area
C-CT: Cardiac computer tomography
DSE: Dobutamine stress echocardiography
LV: left ventricular
LVMi: Left ventricular mass index
RWT: Relative wall thickness
SVi: Stroke volume indexed
Vmax: Aortic valve peak velocity

## Acknowledgements

OPEN, Open Patient data Explorative Network, Odense University Hospital, Region of Southern Denmark.

## Disclosure

None.

### Central Illustration

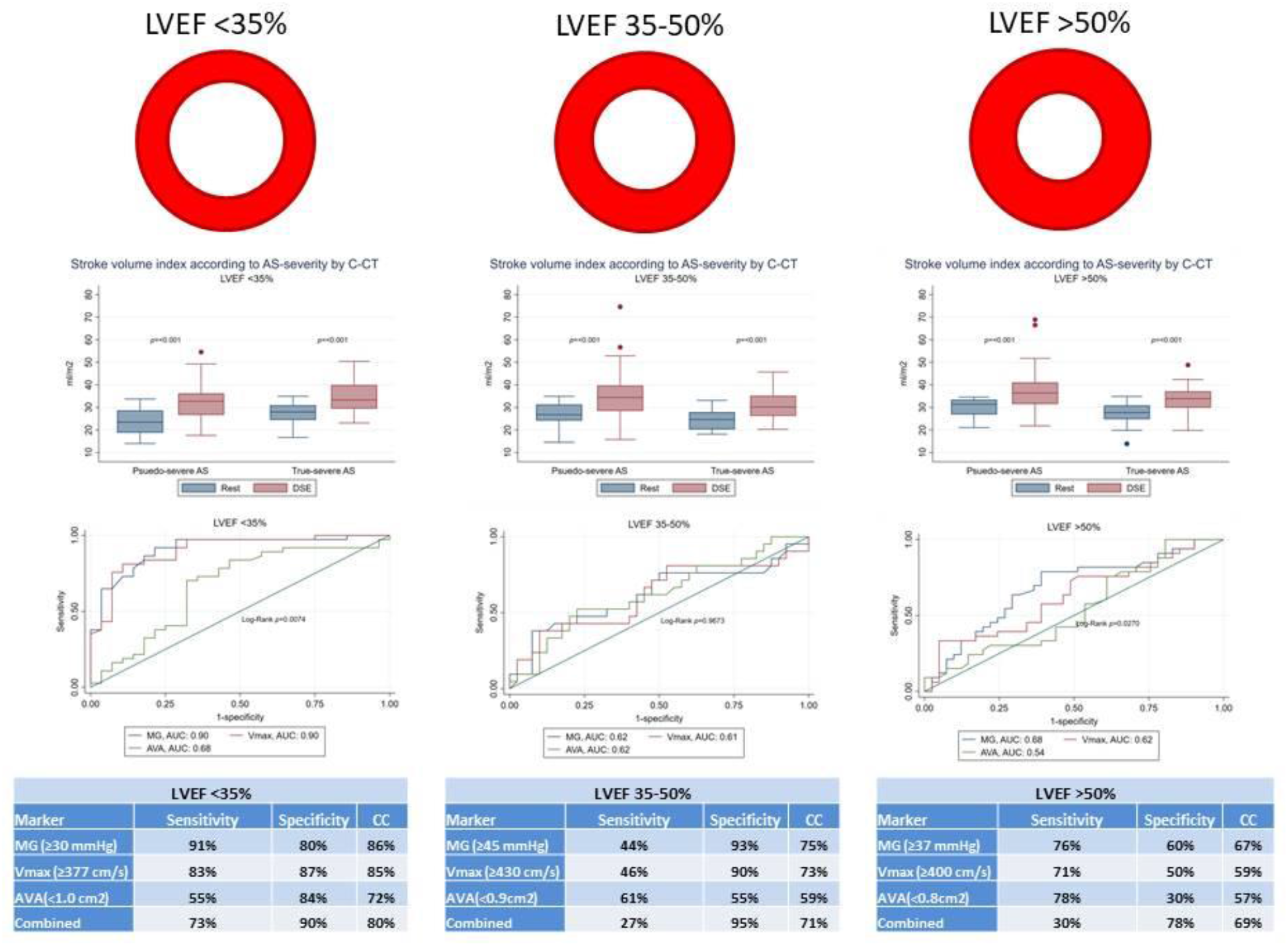

